# Spatial and Climatic Analysis of the 2025 Chikungunya Re-emergence in Mauritius

**DOI:** 10.64898/2026.09.06.26361702

**Authors:** Smita Goorah, Manta Nowbuth, Mahendra Gooroochurn

## Abstract

**Background:** Chikungunya re-emerged in Mauritius in 2025 after a 19-year period with no outbreak. As precipitation and temperature play a key role in mosquito proliferation, the climatic conditions prevailing in Mauritius in 2025 may have influenced the emergence and spread of the outbreak. Our aim in this study was to investigate the spatial distribution of the 2025 chikungunya outbreak in Mauritius, and to determine the association between chikungunya incidence and selected predictor variables using Geographic Information Systems (GIS).

**Methods:** The variables considered were mean altitude, mean air temperature and mean precipitation, population density, road density, number of houses near rivers and the relative deprivation index (RDI) per village council area (VCA) and municipal ward (MW). Rainfall and temperature data were obtained from the climate bulletins of the Mauritius Meteorological Services and were also extracted from the ERA5 Monthly Averaged Data on Single levels available from the platform of Copernicus Climate Data Store. The U.S. Geological Survey Earth Explorer platform was used to access elevation data for Mauritius. In the absence of official case data, reported chikungunya cases were reconstructed from multiple publicly accessible media sources. Information was collected contemporaneously as the outbreak evolved during 2025 and was supplemented by searches of archived material. Cases were georeferenced and aggregated to VCAs and MWs. All data was mapped using QGIS version 3.40.13.

**Results:** The main findings were that the 2025 chikungunya outbreak was significantly associated with elevation and precipitation. Temporal analysis demonstrated that rainfall in the preceding month was the strongest climatic predictor of monthly chikungunya cases. Spatial analysis, however, showed that mean precipitation had a significant (*p*-value < 0.001) and negative association with chikungunya incidence, indicating lower disease occurrence in areas of high precipitation. Mean elevation was significantly (*p*-value < 0.001) and negatively associated with chikungunya incidence, indicating lower disease occurrence in higher-altitude areas. Temperature did not have any significant relationship with chikungunya incidence.

**Conclusions:** Rainfall had opposite associations depending on the analytical scale: preceding-month rainfall was positively associated with temporal case variation, whereas spatially averaged precipitation was negatively associated with incidence. Thus, rainfall exerted distinct temporal and spatial influences on chikungunya transmission, while higher elevation was associated with lower chikungunya incidence. These findings highlight the importance of integrating climatic and spatial analyses to improve surveillance

## Introduction

Chikungunya fever re-emerged in Mauritius in 2025 after previous outbreaks in 2005 and 2006. The first locally-acquired case was reported on the 15 March 2025 (Government Information Service, 2025). The island recorded a total of 1583 cases at the end of the outbreak in September 2025 (Boston University Center on Emerging Infectious Diseases, 2026). The start of the outbreak occurred 2 weeks after the tropical cyclone Garance which generated heavy rainfall across Mauritius after marked deficits in January and February 2025. Chikungunya fever was first characterized in 1952-1953 by Lumsden in Tanzania (Lumsden, 1955). The chikungunya virus (CHIKV) was then isolated by Ross, (1956). The CHIKV is an alphavirus consisting of a single-stranded, positive-sense RNA genome. There are three geographically associated genotypes namely the West African, East/Central/South African (ECSA), and Asian genotypes (Volk, 2010).

Considering the regional context, no major outbreaks were reported in any Indian Ocean islands in the period 2006-2024. In August 2024, there was a widespread resurgence of chikungunya in the neighbouring island of Reunion (WHO, 2025). A study by Frumence *et al*., (2026) showed that precipitation and temperature were significantly associated with the progression of the epidemic in Reunion, both with and without a one-month lag period. The infection subsequently spread to Mauritius. Regional outbreaks highlight the vulnerability of interconnected islands to the spread of arboviral diseases through human movement as CHIKV is typically introduced by infected individuals arriving from epidemic areas. Once introduced in a country, local transmission of the virus can become established where competent vectors are present. *Aedes albopictus*, the principal mosquito vector in both Reunion and Mauritius, proliferates under favourable climatic conditions.

As precipitation and temperature play a key role in mosquito proliferation, the climatic conditions prevailing in Mauritius in 2025 may have influenced the emergence and spread of the outbreak. Island-wide temperatures were consistently above long-term mean (LTM) throughout the year. Our aim in this study was to investigate the spatial distribution of the outbreak and to determine the association between chikungunya incidence and selected predictor variables using Geographic Information Systems (GIS). The variables considered were mean altitude, mean air temperature, mean precipitation, population density, road density, number of houses near rivers and the relative deprivation index (RDI).

## Methods

### Monthly data

Monthly chikungunya case numbers were obtained from official publications of the Ministry of Health and Wellness (2025). Monthly rainfall and mean temperature data were obtained from the climate bulletins of the Mauritius Meteorological Services (2025). Monthly temperature LTMs were obtained from Statistics Mauritius (2025). The chikungunya monthly cases were log-transformed. A multiple linear regression analysis was carried out to test associations between log-transformed chikungunya cases and 1. monthly rainfall and monthly mean temperature; 2. one-month lagged rainfall and one-month lagged mean temperature; 3. one-month lagged rainfall and monthly mean temperature. All data was analysed using Microsoft Excel (Microsoft Corporation, 2021).

### Spatial distribution of chikungunya

In the absence of official case data, reported chikungunya cases were reconstructed from multiple publicly accessible sources, including news media, social media and crowdsourced information. Information was collected contemporaneously as the outbreak evolved during 2025 and was supplemented by searches of archived material. Cases were georeferenced and aggregated to village council areas (VCAs) and municipal wards (MWs). All data was mapped using QGIS version 3.40.13. Chikungunya incidence (per 10,000 of population) was then calculated. The limitations associated with the use of publicly sourced outbreak information from different sources, including potential under-reporting, duplicate reporting and incomplete geographical information, were considered in interpretation of the findings. The data should be considered as best approximation.

### Distribution of elevation

The U.S. Geological Survey (USGS, 2026) Earth Explorer platform was used to access elevation data for Mauritius. The USGS platform distributes elevation data obtained from the NASA Shuttle Radar Topography Mission (SRTM) Global 1 Arc-Second Digital Elevation Model (NASA JPL, 2013). The SRTM data has global coverage and is well validated (Farr et al., 2007).

### Distribution of temperature and precipitation

Temperature and precipitation data for Mauritius were extracted from the ERA5 Monthly Averaged Data on Single levels. It is a reanalysis data which reconstructs the most probable atmospheric conditions based on weather station observations, satellite observations, aircraft, ship and buoy observations as well as weather forecast models (Hersbach et al., 2023). The ERA5 data is freely available from the download platform of Copernicus Climate Data Store (Copernicus Climate Change Service (C3S), ERA5 dataset, 2026).

### Distribution of other predictors

Descriptions of the methodology and the spatial distribution of population density, the number of houses near rivers, and the RDI are available in our previously published study (Goorah et al., 2025). Road network data was obtained from the Mauritius Roads (OpenStreetMap Export) dataset. This free dataset is provided by OpenStreetMap contributors and is distributed through the Humanitarian Data Exchange (OpenStreetMap contributors, 2020).

### Ethics statement

This study used aggregated and estimated publicly available information and did not involve the collection or analysis of identifiable individual-level patient data.

### Statistical analysis

A multiple linear regression was calculated to predict the outcome variable which was the log (chikungunya incidence per VCA and MW) based on the predictor variables as described above. When predictors were highly correlated as observed in a correlation matrix which was carried out, the predictor with the stronger individual correlation with the outcome variable was retained, and a second multiple linear regression was carried out with the reduced predictors.

## Results

Results of the monthly chikungunya cases, monthly mean temperature and total monthly rainfall for the year 2025 in Mauritius are shown in Table 1.

**Table 1:** Monthly chikungunya cases, monthly mean temperature and total monthly rainfall in Mauritius for the period January–December 2025.

| Month in 2025 | Total monthly Rainfall /mm | % Rainfall LTM | Monthly mean temperature LTM/°C (1991-2020) | Monthly mean temperature deviation from LTM /°C | Monthly mean temperature /°C | Monthly chikungunya cases |
| --- | --- | --- | --- | --- | --- | --- |
| Jan | 85 | 30 | 26.3 | 0.8 | 27.1 | 0 |
| Feb | 190 | 59 | 26.4 | 1.15 | 27.55 | 0 |
| Mar | 168 | 57 | 26.0 | 1.2 | 27.2 | 17 |
| Apr | 251 | 122 | 25.1 | 1.18 | 26.28 | 100 |
| May | 286 | 193 | 23.4 | 1.11 | 24.51 | 688 |
| Jun | 147 | 126 | 21.7 | 0.43 | 22.13 | 558 |
| Jul | 149 | 113 | 20.9 | 0.91 | 21.81 | 185 |
| Aug | 136 | 126 | 20.9 | 0.43 | 21.33 | 30 |
| **Sep** | 66 | 78 | 21.5 | 0.5 | 22 | 5 |
| **Oct** | 35 | 48 | 22.7 | 1.04 | 23.74 | 0 |
| **Nov** | 140 | 165 | 24.1 | 0.98 | 25.08 | 0 |
| **Dec** | 171 | 104 | 25.6 | 0.25 | 25.85 | 2 |

A comparison of three multiple linear regression models is shown in Table 2. Results show that one-month lagged rainfall and current temperature has the highest predictive value to log-transformed chikungunya cases.

**Table 2:** Comparison of three multiple linear regression models.

| **Model** | **Rainfall** | **Temperature** | **R<sup>2</sup>** | ***p-value*** |
| --- | --- | --- | --- | --- |
| Both current month | Current | Current | 0.717 | 0.0034 |
| Both lagged | 1-month lag | 1-month lag | 0.762 | 0.0016 |
| Lagged rainfall + current temperature | 1-month lag | Current | 0.797 | 0.0008 |

The multiple linear regression model was statistically significant explaining 79.7% of the variation in monthly chikungunya cases. One-month lagged rainfall was a significant positive predictor of chikungunya cases (*p* = 0.001), indicating that increased rainfall in the preceding month was associated with higher case numbers as shown in Table 3.

**Table 3:**
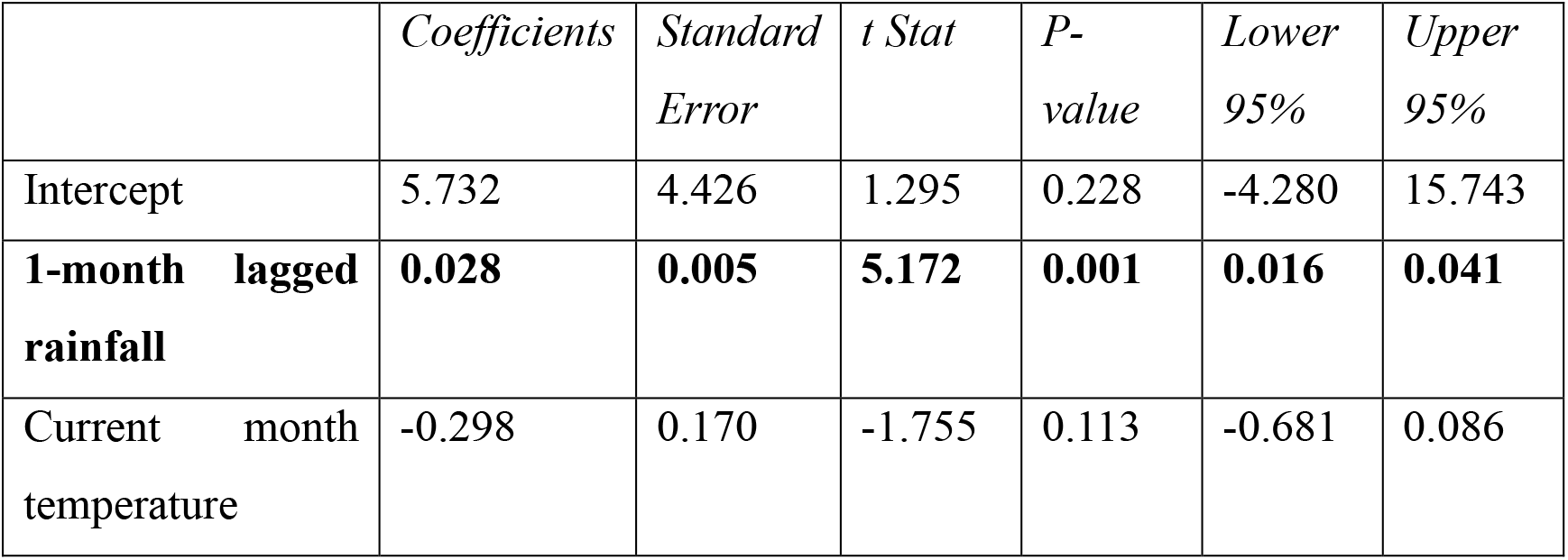
Multiple linear regression coefficients for climatic predictors of log-transformed chikungunya cases in Mauritius, 2025.

**Figure 1:**
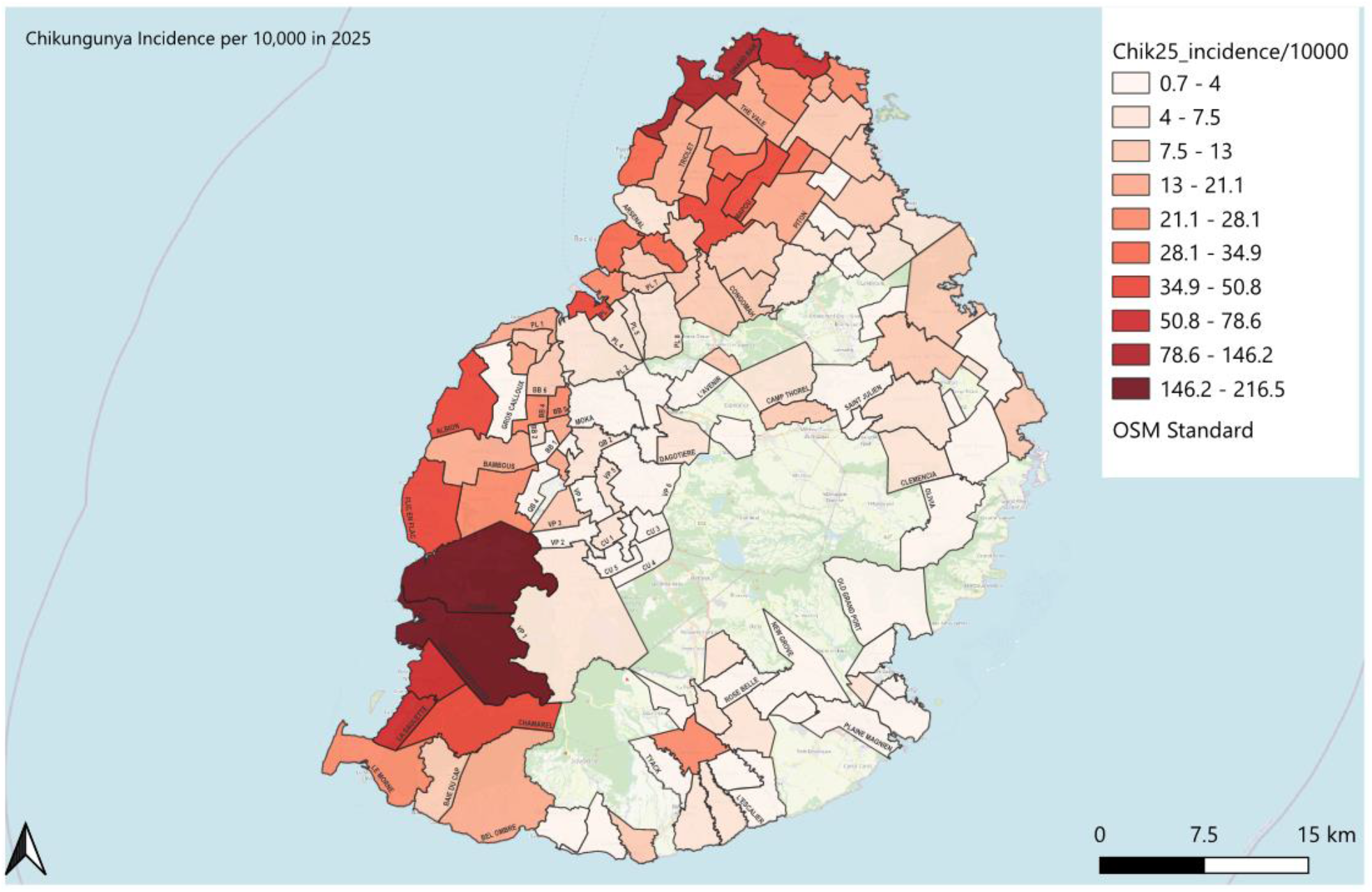
Estimated spatial distribution of chikungunya incidence in Mauritius, 2025. Note: The spatial distribution shown is an estimate based on available publicly reported data and should not be interpreted as the actual official distribution of chikungunya cases.

**Figure 2:**
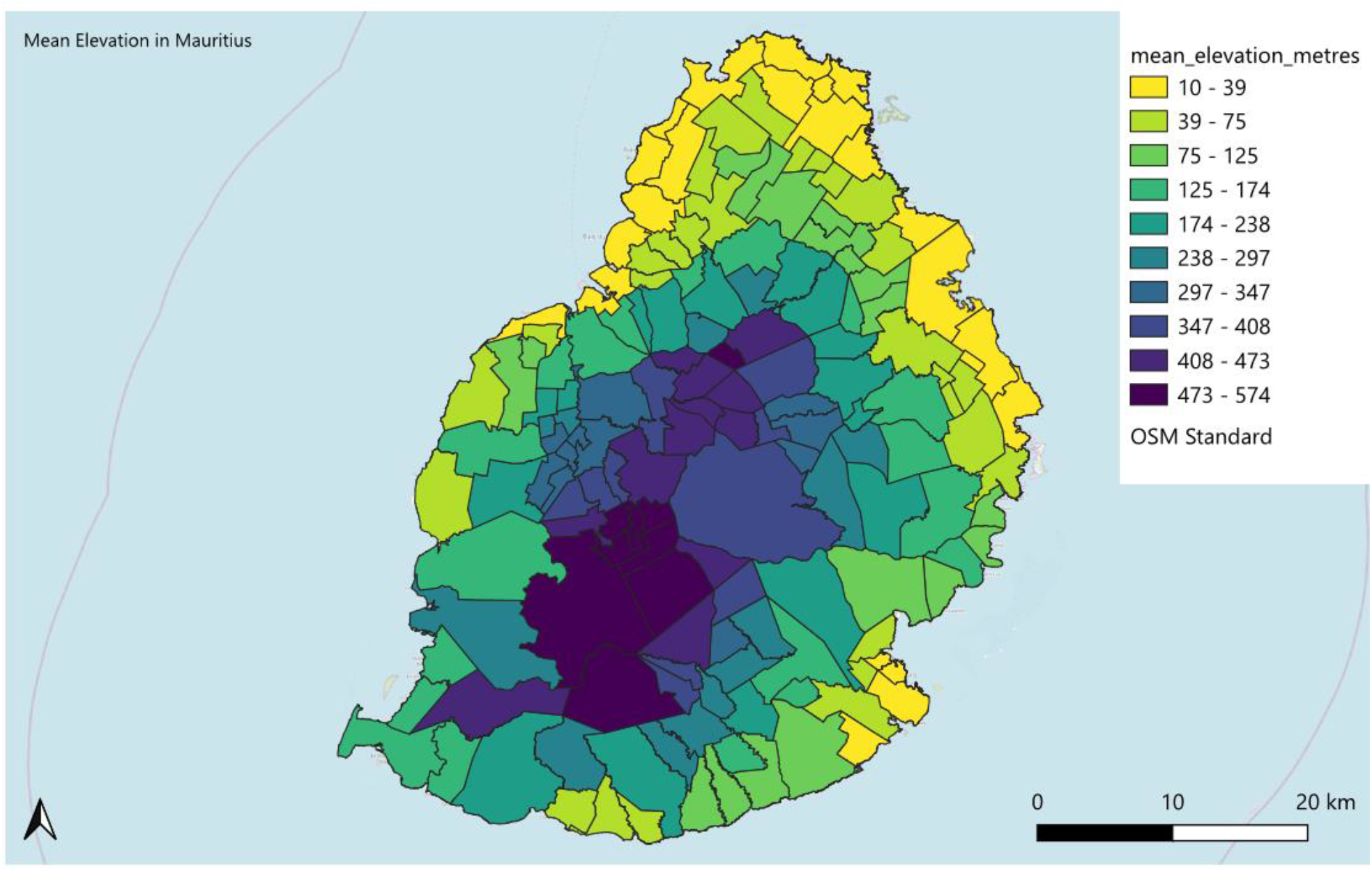
Spatial distribution of mean elevation by MW and VCA

**Figure 3:**
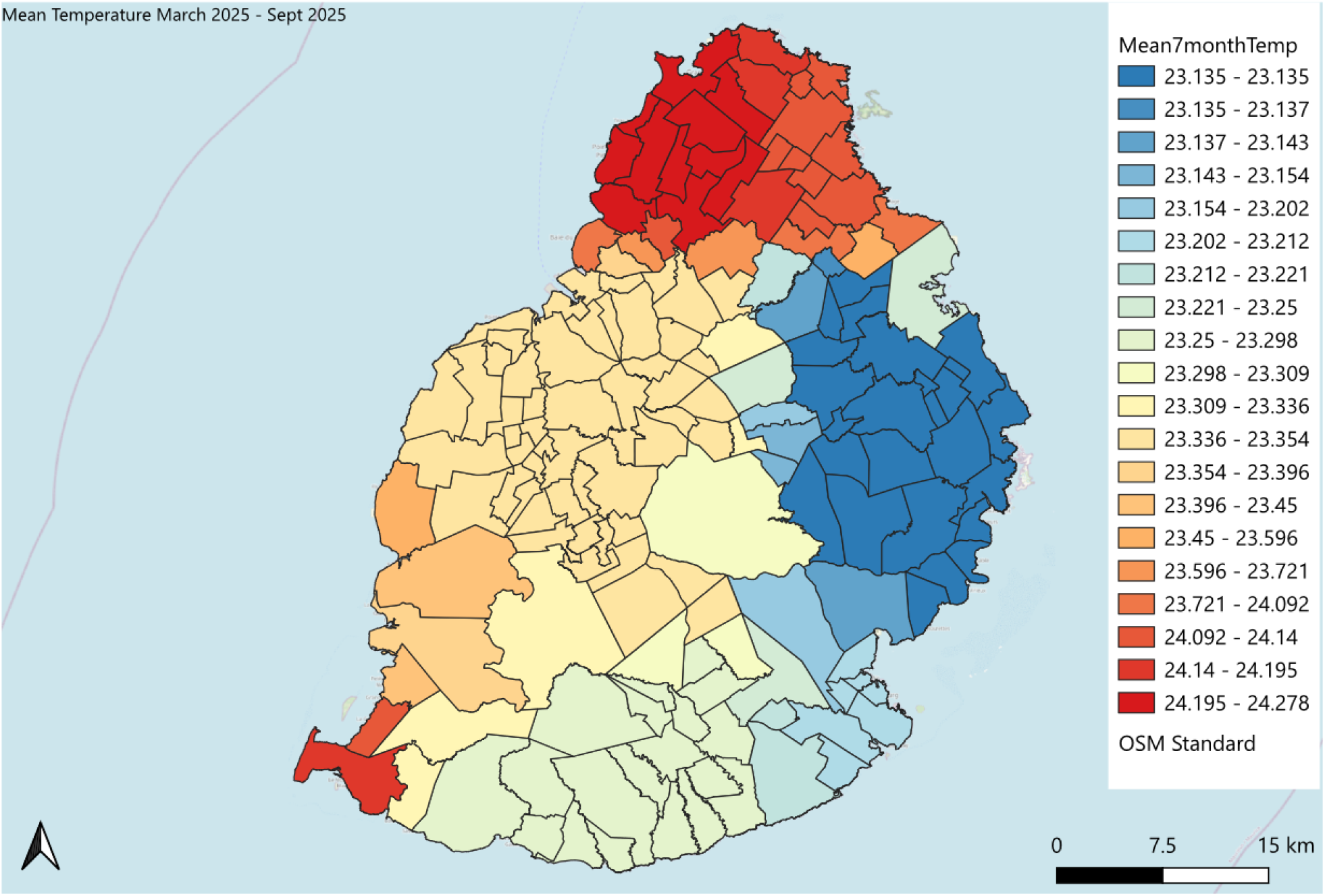
Spatial distribution of mean 7-month temperature by MW and VCA

**Figure 4:**
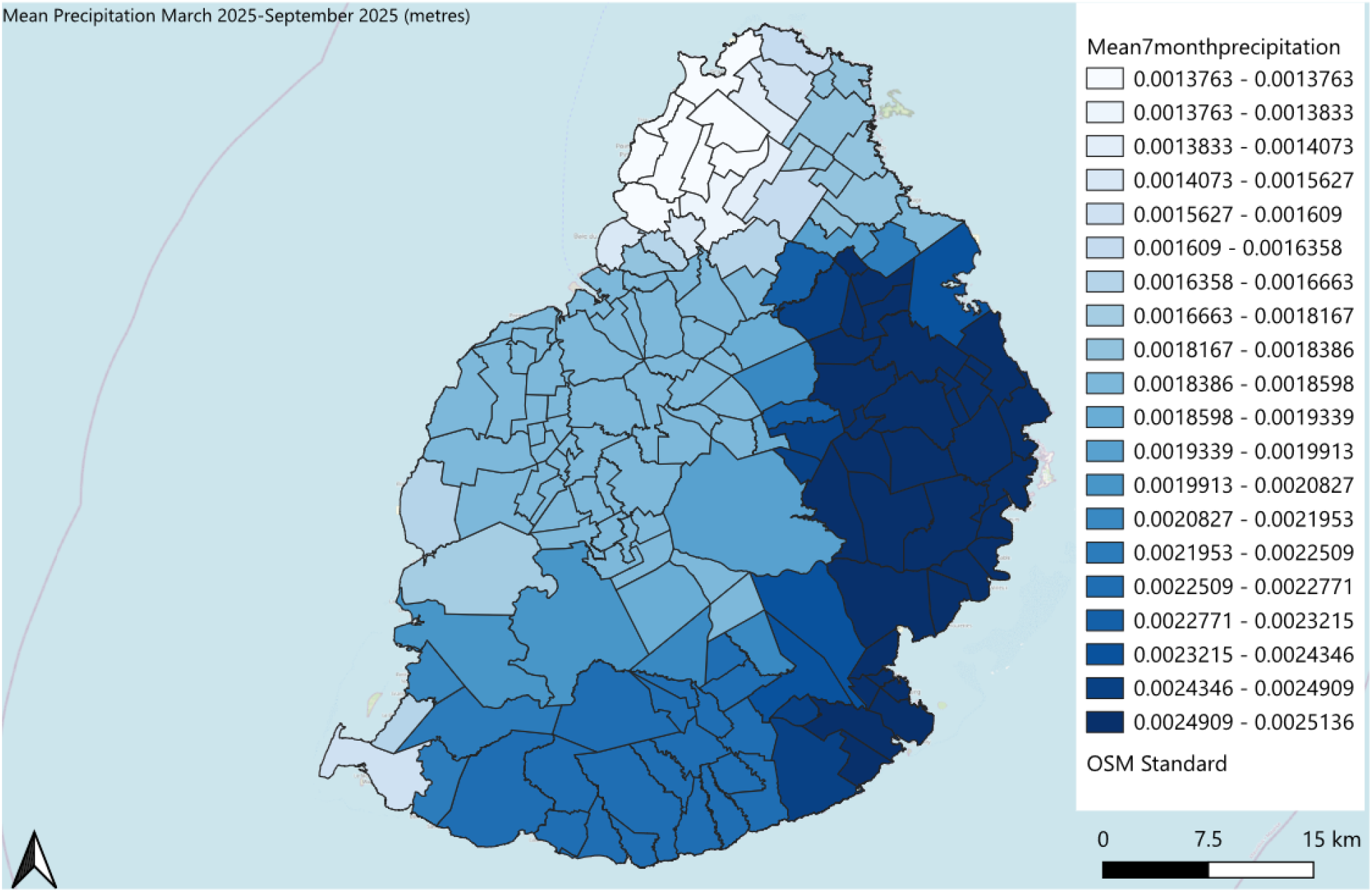
Spatial distribution of mean 7-month precipitation by MW and VCA

On statistical analysis using the seven predictors, the highest correlation was road density vs population density with a strong positive correlation of 0.88. This could be explained by the fact that VCAs and MWs with high population density tended to have high road density. Road density was retained as a predictor in the final model while population density was removed.

There was a strong negative correlation of −0.77 between mean temperature vs mean precipitation. indicating that cooler areas tended to have higher rainfall, while warmer areas tended to have lower rainfall. Mean precipitation was retained as a predictor in the final model while mean temperature was removed.

The following model as shown in Table 4 was obtained after removal of the two highly correlated variables: population density and mean temperature.

**Table 4:**
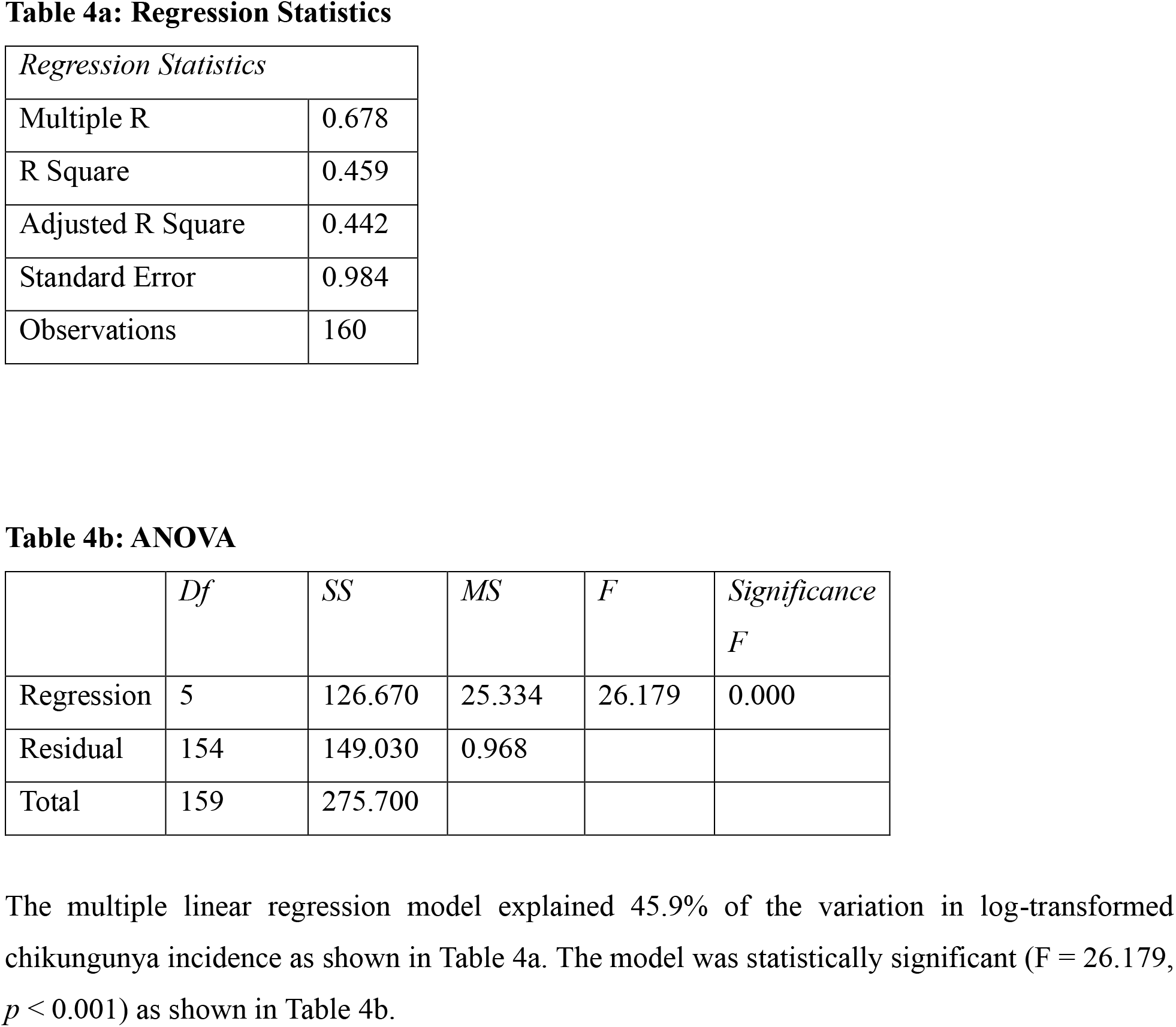
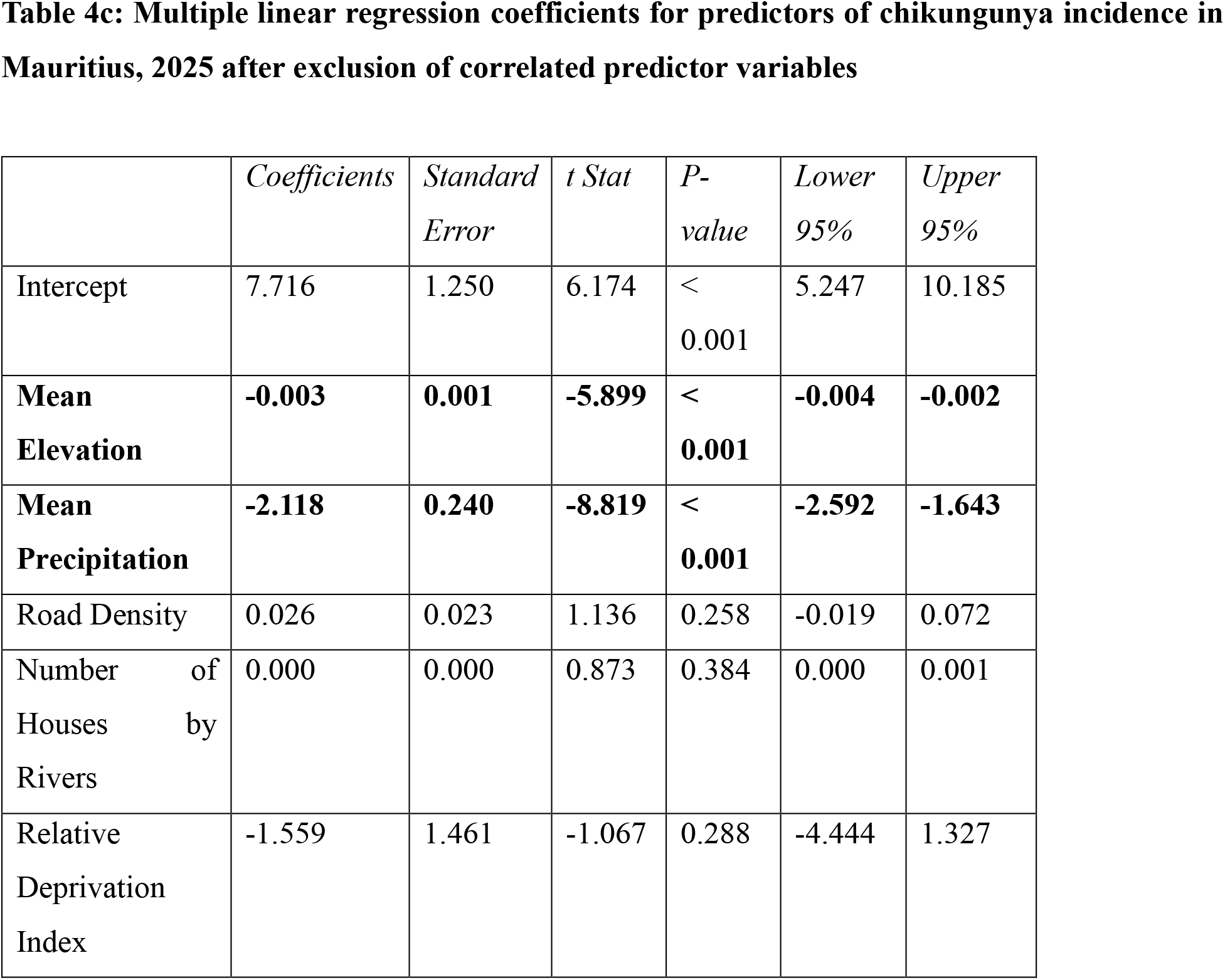
Final multiple linear regression analysis of factors associated with chikungunya incidence after removal of correlated predictor variables.

Results from the multiple regression analysis showed that mean elevation (*p* < 0.001) and mean precipitation (*p* < 0.001) were significant predictors after removal of the highly correlated variables.

Mean elevation was significantly (*p*-value < 0.001) and negatively associated (coefficient= −0.003) with chikungunya incidence, indicating lower disease occurrence in higher-altitude areas.

Mean precipitation showed a significant (*p*-value < 0.001) and negative association with a coefficient of −2.118 with chikungunya incidence, suggesting that MWs and VCAs with higher precipitation experienced lower disease incidence during the study period.

In this model, road density, the number of houses near rivers and RDI were not significantly associated with chikungunya incidence (*p* > 0.05).

## Discussion

The main findings of this study were that the 2025 chikungunya outbreak was significantly associated with elevation and precipitation. The analysis of monthly climatic variables in relation to chikungunya cases indicated that total rainfall in the preceding month was the strongest climatic predictor. The significant positive association with one-month lagged rainfall was a measure of temporal variability, and was consistent with the time required for the aquatic phases of the life cycle of the mosquito to occur. On the other hand, temperature did not have any significant association with chikungunya cases.

Spatial mapping and analysis showed that mean elevation and mean 7-month precipitation per MW and VCA were identified as significant negative predictors chikungunya incidence in Mauritius in 2025. Thus, we note that precipitation had a positive temporal effect and a negative spatial effect. Areas with the highest average precipitation did not have the highest transmission of chikungunya. This result could be explained by the flushing out of mosquito breeding sites in excessive rainfall events. Indeed, the Central Plateau, East and South of the island had intense rainfall events which may explain a lower incidence of chikungunya. On the other hand, the dry localities in the West and North experienced high incidences of chikungunya especially in the coastal areas. As the minimum average precipitation was 1.38 mm per day across the island, it is plausible that there was sufficient rainfall for mosquito breeding even in the driest regions.

The mean monthly temperature and the mean temperature during March–September 2025 were not significantly associated with chikungunya incidence. Throughout the study period, monthly temperatures remained consistently above the LTM, while mean temperatures across MWs and VCAs varied only between 23.1°C to 24.3°C. This narrow range lies within the favourable thermal conditions for the *Ae. albopictus* mosquito development, survival and biting frequency as well as CHIKV replication within the mosquito. Hence, our study shows that temperature was not the primary factor explaining the temporal or spatial variation in chikungunya incidence during the outbreak.

### Elevation

Elevation was a significant negative predictor of chikungunya incidence in 2025. Possible explanations are that higher altitudes are cooler with more wind exposure, and these do not favour mosquito survival or virus replication. In addition, land use patterns and housing characteristics may differ in higher elevations areas. Human behaviour may also contribute to reduce human-mosquito interactions. For example, people may wear long-sleeved clothes, remain indoors and close doors and windows more commonly in cooler, windier and more humid areas. Whereas, inhabitants of coastal regions may wear lighter clothing, spend more time outdoors and tend to leave windows open, therefore increasing opportunities for mosquito bites.

### Other possible drivers

The final regression model explained 45.9% of the variation in log-transformed chikungunya incidence. Thus, approximately 54% of the variation remained unexplained and may be attributable to other factors. Variables such as behavioural patterns, use of mosquito repellents, vector control interventions, housing characteristics, relative humidity and previous immunity may have contributed to variations in incidence. In addition, differences in healthcare-seeking behaviour, case reporting, and the spatial resolution of climatic data may have introduced further unexplained variability.

## Conclusion

Mauritius experienced a re-emergence of chikungunya in 2025. Spatial analysis identified altitude and mean precipitation as significant negative predictors of chikungunya incidence in our study. In addition, temporal analysis demonstrated that rainfall in the preceding month was the strongest climatic predictor of monthly chikungunya cases, consistent with the delayed effect of rainfall on mosquito breeding and subsequent virus transmission. These findings demonstrate that precipitation exerts distinct temporal and spatial influences on chikungunya transmission and highlight the importance of integrating climatic and spatial analyses to improve surveillance, risk assessment and preparedness for future arboviral outbreaks in Mauritius.

## Data Availability

All data produced in the present work are contained in the manuscript.

## References

BOSTON UNIVERSITY CENTER ON EMERGING INFECTIOUS DISEASES, 2026. BEACON: Biothreats Emergence, Analysis and Communications Network report.

COPERNICUS CLIMATE CHANGE SERVICE, 2023. ERA5 monthly averaged data on single levels from 1940 to present. Copernicus Climate Change Service (C3S) Climate Data Store (CDS). DOI: 10.24381/cds.f17050d7

[Attribution-Copernicus programme: Generated using or contains modified Copernicus Climate Change Service information 2026. Neither the European Commission nor ECMWF is responsible for any use that may be made of the Copernicus information or data it contains]

Farr, T.G., Rosen, P.A., Caro, E., Crippen, R., Duren, R., Hensley, S., Kobrick, M., Paller, M., Rodriguez, E., Roth, L., Seal, D., Shaffer, S., Shimada, J., Umland, J., Werner, M., Oskin, M., Burbank, D., Alsdorf, D., 2007. The Shuttle Radar Topography Mission. Reviews of Geophysics, 45(2), RG2004. DOI: 10.1029/2005RG000183.

Frumence, E., Klitting, R., Serres, K., Shao, Y., Monti, F., Vincent, M., Gill, M.S., Suchard, M.A., Lemey, P., De Lamballerie, X., Jaffar-Bandjee, M.-C. Dellicour S., 2026. Unravelling the epidemiological and dispersal dynamics of the 2024–2025 chikungunya virus epidemic on Réunion island. Preprint. DOI: 10.64898/2026.01.07.26343606

Goorah, S., Nowbuth, M., Gooroochurn, M., 2025. Identifying drivers of dengue fever outbreaks in Mauritius using Geographic Information System. Jàmbá: Journal of Disaster Risk Studies, 17(2), a1740. DOI: 10.4102/jamba.v17i2.1740.

GOVERNMENT INFORMATION SERVICE, 2025. Health authorities call on public to collaborate after detection of local Chikungunya case. Government of Mauritius, 18 March 2025.

Hersbach, H., Bell, B., Berrisford, P., Biavati, G., HorÁNyi, A., MuÑOz Sabater, J., Nicolas, J., Peubey, C., Radu, R., Rozum, I., Schepers, D., Simmons, A., Soci, C., Dee, D., ThÉPaut, J.-N., 2023. ERA5 monthly averaged data on single levels from 1940 to present. Copernicus Climate Change Service (C3S) Climate Data Store (CDS). DOI: 10.24381/cds.f17050d7.

Lumsden W.H., 1955. An epidemic of virus disease in Southern Province, Tanganyika Territory, in 1952–53. II. General description and epidemiology. Transactions of the Royal Society of Tropical Medicine and Hygiene, 49(1):33–57.

MAURITIUS METEOROLOGICAL SERVICES, 2025. Climate Bulletins. Vacoas: Mauritius Meteorological Services.

MICROSOFT CORPORATION, 2021. Microsoft Excel. Version 2021. Redmond, WA: Microsoft Corporation.

MINISTRY OF HEALTH AND WELLNESS, 2025. Annual report on performance for financial year 2024–2025. Port Louis: Ministry of Health and Wellness.

NASA JET PROPULSION LABORATORY (JPL), 2013. NASA Shuttle Radar Topography Mission (SRTM) Global 1 Arc Second. Distributed by the U.S. Geological Survey Earth Resources Observation and Science (EROS) Center.

OPENSTREETMAP CONTRIBUTORS, 2020. Mauritius Roads (OpenStreetMap Export). Humanitarian Data Exchange. [dataset]. Available from: https://data.humdata.org/dataset/a3923cd8-88d1-461a-b1fe-c133dd25e115

Ross, R.W., 1956. The Newala epidemic. III. The virus: isolation, pathogenic properties and relationship to the epidemic. The Journal of Hygiene, 54:177–191.

STATISTICS MAURITIUS, 2025. Environment Statistics – Year 2024. Economic and Social Indicators, Issue No. 1873. Port Louis: Statistics Mauritius.

U.S. GEOLOGICAL SURVEY (USGS), 2026. SRTM 1 Arc-Second Global [Digital elevation model]. Available at: https://earthexplorer.usgs.gov.

Volk, S.M., Chen, R., Tsetsarkin, K.A., Adams, A.P., Garcia, T.I., Sall, A.A., Nasar, F., Schuh, A.J., Holmes, E.C., Higgs, S., Maharaj, P.D., Brault, A.C., Weaver, S.C., 2010. Genome-scale phylogenetic analyses of chikungunya virus reveal independent emergences of recent epidemics and various evolutionary rates. Journal of Virology, 84(13), 6497–6504.

WORLD HEALTH ORGANIZATION, 2025. Chikungunya virus disease – Global situation. Disease Outbreak News, 3 October.

